# COVID-19 projections for reopening Connecticut

**DOI:** 10.1101/2020.06.16.20126425

**Authors:** Forrest W. Crawford, Zehang Richard Li, Olga Morozova

**Affiliations:** Department of Biostatistics, Yale School of Public Health; Department of Statistics & Data Science, Yale University; Department of Ecology & Evolutionary Biology, Yale University; Yale School of Management

## Abstract

**Key Points:**

- Closure of schools and the statewide “Stay Safe, Stay Home” order have effectively reduced COVID-19 transmission in Connecticut, with model projections estimating incidence at about 1,300 new infections per day.
- If close interpersonal contact increases quickly in Connecticut following reopening on May 20, the state is at risk of a substantial increase of COVID-19 infections, hospitalizations, and deaths by late Summer 2020.
- Real-time metrics including case counts, hospitalizations, and deaths may fail to provide enough advance warning to avoid resurgence.
- Substantial uncertainty remains in our knowledge of cumulative COVID-19 incidence, the proportion of infected individuals who are asymptomatic, infectiousness of children, the effects of testing and contact tracing on isolation of infected individuals, and how contact patterns may change following reopening.

## Introduction

Connecticut is one of the US states most severely impacted by the COVID-19 pandemic, with over 37,000 cases, 11,000 hospitalizations, and 3,400 deaths [1, 2]. On March 8, 2020, the first Connecticut COVID-19 case was identified, and the state began interventions to slow transmission in mid March 2020. On March 17, Governor Ned Lamont ordered all in-person classes at K-12 schools canceled [3], and later extended the closure for the remainder of the 2019-2020 academic year [4–6]. The Governor issued a statewide “Stay Safe, Stay Home” order to take effect on March 23 [7]. Evidence from device mobility data suggests that Connecticut residents reduced travel outside the home, and increased the number of hours per day spent at home [8, 9]. At the same time, the Governor ordered businesses closed, with the exception of essential businesses that could remain open with additional restrictions and guidelines to minimize close contact and risk of transmission.

The number of hospitalized COVID-19 patients in Connecticut peaked in mid April, and began a slow decline. In early May, Governor Lamont issued plans and guidance for reopening the state, a process that began on May 20. The Connecticut Department of Economic and Community Development issued rules and guidelines for opening particular business sectors [10]. Monitoring COVID-19 test results, expanding access to testing, and contact tracing are part of the state’s intervention strategy. Contact tracing will be implemented using the state’s *ContaCT* platform [11], and the state publishes daily testing reports [e.g. 12]. The Governor announced goals to scale up COVID-19 viral testing to 42,000 tests per day [13, 14], but testing capacity is still limited [15].

As Connecticut reopens, policymakers need access to reliable information about both the current state of the pandemic and likely future outcomes. Elected officials and public health agencies already have access to near real-time information about COVID-19 testing, case counts, hospitalizations, and deaths. However, these data may not provide timely insight into the current and future dynamics of COVID-19 transmission. Because viral testing of asymptomatic individuals is not yet widespread, symptomatic individuals at least 4-5 days from infection may constitute the majority who seek testing [16–22], and COVID-19 case counts or positive test proportion may be poor surrogate measures of current infection prevalence. Hospitalization may not occur until more than 2 or 3 weeks following infection, and death may not occur for weeks after that [17, 23–27]. Furthermore, current data may not provide information about what is likely to occur under the Governor’s stated reopening plans or interventions to be implemented in the future. As the state scales up its testing capacity, expands contact tracing, and initiates phased reopening of businesses, historical data on cases, hospitalizations, and deaths may not give insight into the possible effects of these interventions on future incidence. Finally, early warning systems based on these lagging metrics may fail to provide state policymakers with the predictive information they need to initiate a reversion to a more restrictive phase or implement other interventions to blunt a coming wave of new infections.

In this report, we use a mathematical infectious disease transmission model to project COVID-19 incidence, hospitalizations, and deaths in the state of Connecticut through August 31, 2020. We consider population-level contact scenarios informed by the Connecticut Governor’s phased reopening plans. Model parameters are calibrated using data on hospitalizations and deaths in Connecticut and estimates from the literature on clinical epidemiology of COVID-19. A separate technical report describes the data, transmission model, and calibration in detail [28]. The primary purpose of this report is to provide state and local decision-makers with information they can use to plan reopening of the state in a way that minimizes the risk of a resurgence in COVID-19 cases, hospitalizations, and deaths. The secondary purpose of this report is to assist state agencies and non-governmental organizations in implementation of public health responses, including testing, contact tracing, and design of incidence, infection prevalence, and seroprevalence studies.

## COVID-19 projections: March 1 to August 31, 2020

We present projections of COVID-19 incidence, hospitalizations, and deaths from March 1 to August 31, 2020. Projections and 95% uncertainty intervals show the range of possible outcomes over the Summer under “slow” and “fast” reopening scenarios. Projections prior to May 20, 2020 are calibrated to historical hospitalization and death data obtained from the Connecticut Department of Public Health [29]. County-level hospitalization and bed capacity are obtained from the Connecticut Hospital Association and CHIMEData [30, 31]. This analysis does not use individual-level patient data. Projections beyond May 20 depend on assumptions about the nature of contact and viral testing in the future, under the Governor’s plan for reopening the state [32].

A separate technical report describes the data, transmission model, calibration, and uncertainty calculation in greater detail [28]. Briefly, we employ a modified compartmental susceptible-exposed-infectious-removed (SEIR) transmission model that accommodates asymptomatic, mildly symptomatic, and severe cases, with additional compartments for hospitalizations, unhospitalized severe infections in the case of hospital overflow, patients in nursing homes and assisted living facilities, and deaths. Exposed individuals are assumed to become infectious 4 days after infection [16–22]. Mildly symptomatic individuals are assumed to self-isolate on average 4 days after they become infectious (or 2.5 days after they develop symptoms) [21, 33, 34], and subsequently remain in isolation until recovery. Asymptomatic individuals are assumed to be less transmissible compared to symptomatic cases, but remain infectious for an average of 7 days in the absence of widespread testing [35]. All severe cases are assumed to require hospitalization. Severe cases occurring in closed facilities, such as nursing homes or assisted living facilities, are tracked separately. Only severe cases can result in death. We assume that COVID-19 infection confers lasting immunity to reinfection at least within the projection time interval of March 1 to August 31, 2020.

In this report we present results assuming 50% of infections are asymptomatic, though there is still uncertainty about this quantity [20, 36–40]. We assume that 10% of symptomatic infections are severe and require hospitalization [17, 21, 23, 41]. Severe cases transition through a latent period, pre-hospitalization infectious period, hospitalization period, and eventually either recover or die. We assume that hospitalizations and deaths are reported with a lag and account for this lag when we match projections to observations. Reporting lags along with average time spent at various stages preceding hospitalization and death are calibrated to observations. Average reporting lag for hospitalizations is shorter than that of deaths due to the latter being a combination of deaths occurring in hospitals and in nursing facilities. Figure 1 shows current COVID-19 hospitalization census and cumulative death projections from March 1 to May 20, 2020. Projections match observed data well, and data points largely fall within prediction uncertainty regions.

**Figure 1:**
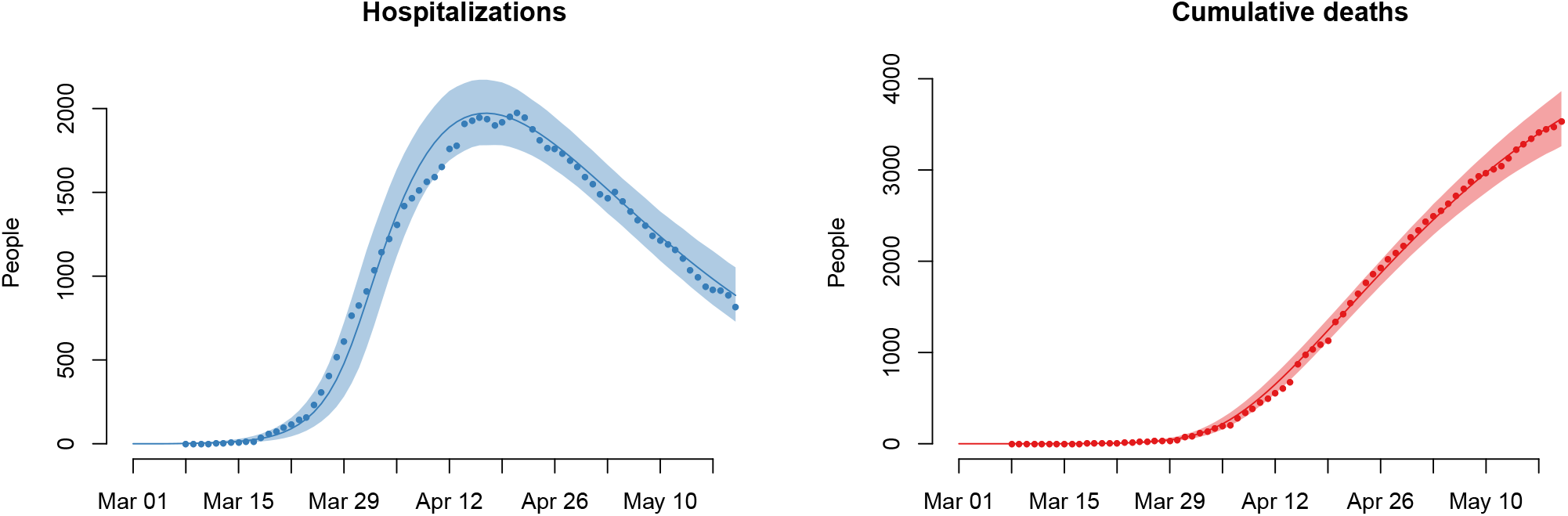
Model projections of current hospitalizations and cumulative deaths, with observed data, from March 1 to May 20, 2020. Solid lines show average model projections and the intervals show 95% uncertainty regions. Observed data are overlaid as solid dots. The figure is reproduced from [28], doi: https://doi.org/10.1101/2020.06.12.20126391.

Additional calibrated projections under low (36%) and high (70%) asymptomatic fractions are provided in a separate technical report [28]. Briefly, when asymptomatic fraction is high, we see more infections, hospitalizations and deaths by the end of Summer. Under slow reopening, hospitalization capacity is not expected to be overwhelmed under any of the scenarios. In case of fast reopening, hospitalization capacity would likely be exceeded sooner if the asymptomatic fraction is high. Differing assumptions about the asymptomatic fraction can result in large differences in estimated cumulative incidence proportion. By the end of Summer, cumulative incidence may be as low as 0.1 under low asymptomatic fraction with slow reopening, or as high as 0.6 under high asymptomatic fraction with fast reopening. The latter scenario predicts the second peak of infections to occur in mid-August, while all other scenarios predict that the second peak would not occur until later in the Fall. While a higher asymptomatic fraction would result in a higher number of deaths in the short-term, the total number of deaths by the end of the epidemic would likely be higher if the asymptomatic fraction is low.

Though we present projections for the state of Connecticut as a whole, the COVID-19 transmission model is structured by county, with transmission allowed between adjacent counties. Reopening scenarios are represented in the projection model by a schedule of proportional increases in contact, beginning on the stated reopening date of May 20, 2020. Slow versus fast reopening scenarios are parameterized by monthly versus biweekly release of 10% of suppressed contact. We estimate that the combined effect of school closure and the “Stay Safe, Stay Home” order reduced contact by about 85% below baseline. Following reopening, viral testing and contact tracing are assumed to increase the rate of isolation of infected individuals. In particular, we assume that asymptomatic individuals are infectious for an average of 7 days, and mildly symptomatic cases are infectious for 4 days, of which 1.5 days is assumed to constitute a pre-symptomatic infectious period [21, 22, 33–35]. Expanded testing and contact tracing, implemented in the weeks following reopening, are assumed to reduce the average duration of infectiousness of asymptomatic individuals to 5.8 days and mildly symptomatic to 2.7 days.

### Projections under slow reopening

Figure 2 shows projections of daily incidence, hospitalizations census, and cumulative deaths under a “slow” reopening scenario. At one-month intervals, 10% of the suppressed contact (relative to March 1 baseline) is released. In these projections, contact rates are at baseline in early March, dropping an estimated 85% of contacts due to closure of schools and the “Stay Safe, Stay Home” order. Under this slow reopening scenario, daily incidence increases following reopening, but decays and stays low until mid August. Hospital census continues its gradual decline through June, flattening in July, and rising again slightly in August. Deaths rise slowly, with the greatest increases occurring in August. Between 4,400 and 6,800 deaths are projected by September 1.

**Figure 2:**
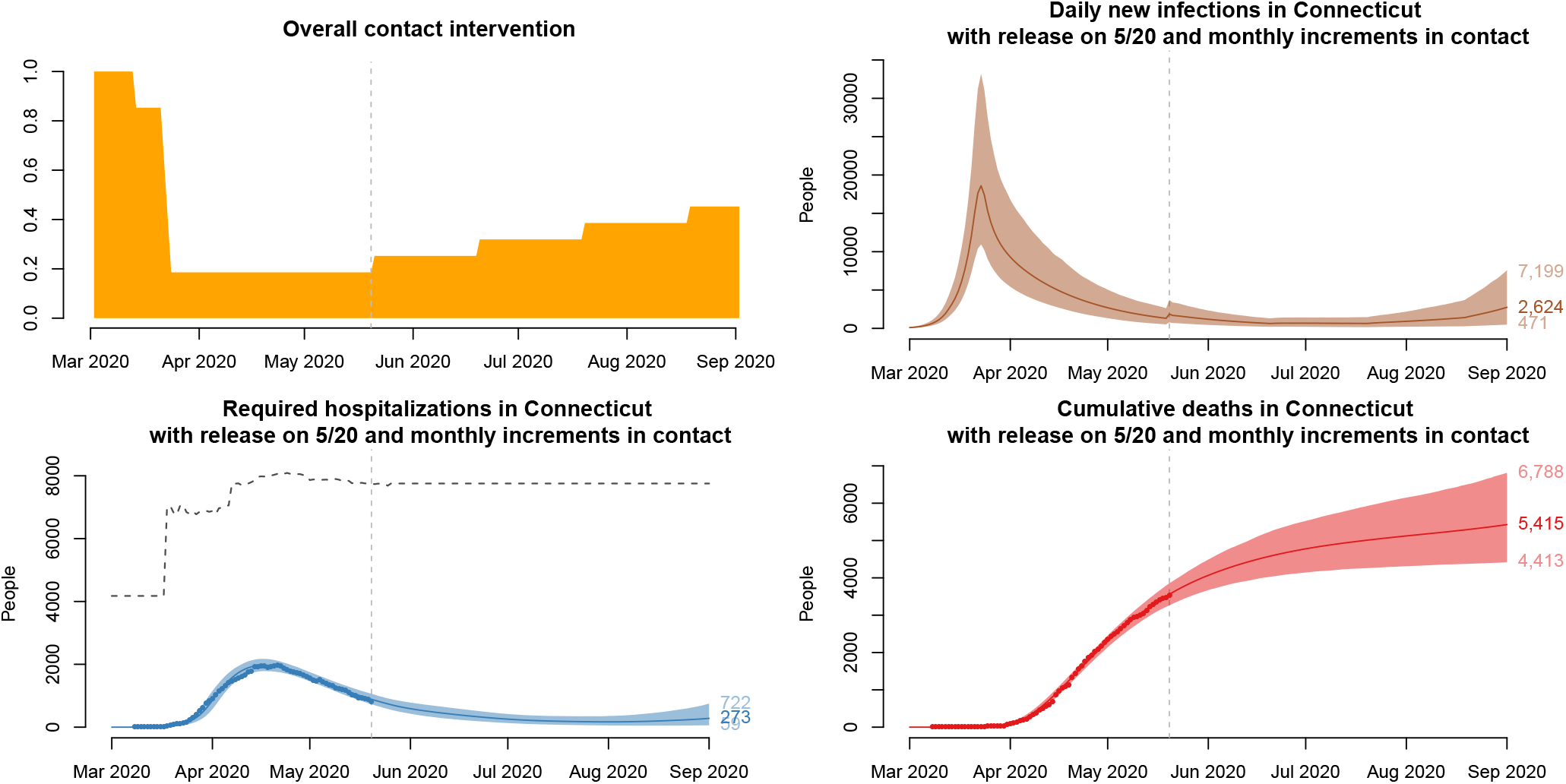
Projections under a “slow” reopening scenario in which 10% of suppressed contact is released every 30 days, starting on May 20, 2020, with 95% uncertainty intervals. Clockwise from top left: population-level historical and projected contact patterns, daily COVID-19 incidence, hospital census, and deaths. The dashed line above hospitalization projections is an estimate of the hospital bed capacity in Connecticut [30]. The figure is reproduced from [28], doi: https://doi.org/10.1101/2020.06.12.20126391.

### Projections under fast reopening

Figure 3 shows projections of daily incidence, hospitalizations census, and cumulative deaths under a “fast” reopening scenario. At two-week intervals, 10% of the suppressed contact is released. In contrast to the “slow” scenario described above, here contact increases rapidly, and by the end of July, daily incidence rises above levels seen in March. By the beginning of September, hospital capacity may be exceeded, resulting in a sharp rise in deaths. Though there is uncertainty about the speed and timing of this predicted rise in hospitalizations and deaths in July-August under a fast reopening scenario, the second wave is likely to occur. Between 5,200 and 12,000 deaths are projected by September 1.

**Figure 3:**
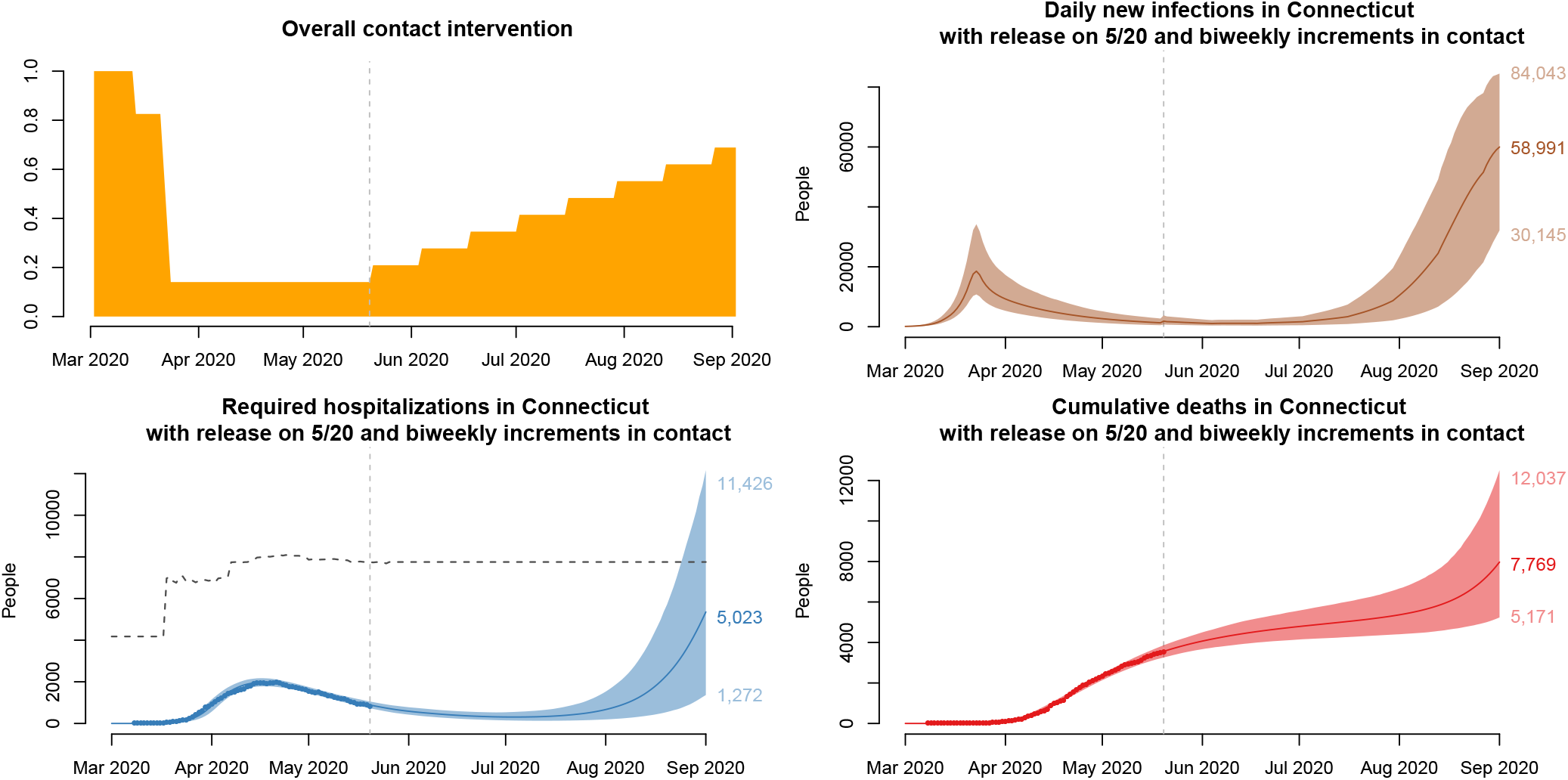
Projections under a “fast” reopening scenario in which 10% of suppressed contact is released every 14 days, starting on May 20, 2020, with 95% uncertainty intervals. Clockwise from top left: population-level historical and projected contact patterns, daily COVID-19 incidence, hospital census, and deaths. The dashed line above hospitalization projections is an estimate of the hospital bed capacity in Connecticut [30], which may be exceeded during August in this scenario. The figure is reproduced from [28], doi: https://doi.org/10.1101/2020.06.12.20126391.

## Implications

The COVID-19 outbreak in Connecticut is currently declining, with hospitalization census decreasing on most days since mid April. Daily incidence at the end of May is estimated to be 1,309 (95% uncertainty interval 505–2,720) new infections per day. However, as the state reopens, there is a risk that increased contact will result in a second wave of COVID-19 transmission, leading to an increase in cases, hospitalizations and deaths. If contact remains low throughout the Summer, with only minimal increases due to reopening, a possible second wave will be blunted and pushed into Fall of 2020. On the other hand, if contact returns quickly to a level seen in early March, then a sharp resurgence in hospitalizations and deaths is likely to occur in August. We do not know with certainty how contact patterns in the Connecticut population will change following reopening on May 20, but it is likely that the true pattern will fall between the slow and fast reopening scenarios described in this report.

### Risk of resurgence following reopening

Projections under a fast reopening scenario suggest a rapid rise in new infections in June and July, followed by an increase in hospitalizations and deaths in August. If transmission increases quickly following reopening, indications of this increase might not be apparent in case counts and hospital data until mid to late July, as the first post-reopen infections become hospitalizations. Therefore it is important for public health authorities to closely monitor new hospitalizations as well as hospital census for signs of increased numbers of COVID-19 cases requiring healthcare. Transmission model projections like the ones presented here may provide more timely warnings than contemporaneous data on cases.

### Importance of testing to detect changes in incidence and isolate infectious individuals

Enhanced viral testing, especially of asymptomatic individuals, may help provide an early warning of increased transmission following reopening. Because there is a roughly 3-week lag between a new infection and possible hospitalization of the infected individual, widespread and frequent testing could detect increased incidence before the first new symptomatic cases present to the hospital. Daily new infections are not observable in real time, but can be estimated using model projections like those presented here. In addition to its usefulness as an early warning system, widespread testing of symptomatic and asymptomatic people, followed by contact tracing, also constitutes a public health intervention. When positive individuals isolate themselves following receipt of a positive test result, they shorten the time during which they expose their contacts to their infection. In this way, testing and contact tracing can hasten isolation of infectious individuals, thereby reducing population-level transmission. In the scenarios presented above, we assume that testing and contact tracing shorten the average infectious period of all asymptomatic individuals from 7 to 5.8 days, and the infectious period of mildly symptomatic individuals from 4 days to 2.7 days. However, widespread testing, contact tracing, and subsequent isolation of individuals who test positive could further reduce the time interval of possible transmission and thereby further suppress incidence.

### Model projections to inform seroprevalence studies

Expanded and accurate antibody testing in Connecticut could dramatically improve knowledge about seroprevalence – the proportion of people in Connecticut who have evidence of prior COVID-19 infection. If this proportion were more precisely known, the fraction of infected individuals who are asymptomatic could be estimated with greater certainty. Currently available data on deaths, hospitalizations and registered cases is consistent with a wide range of asymptomatic infections proportions. This knowledge would reduce uncertainty in future projections because it would reduce uncertainty about the number of people who have already been infected. Approximating cumulative incidence by the cumulative number of registered cases may be seriously misleading, because it omits individuals who experienced asymptomatic infection, or those who were never tested. If infection confers lasting immunity, then information about the true cumulative incidence can give insight into the extent of “herd immunity”, and the effective reproductive number which depends on the fraction of the population that remains susceptible. Knowledge of cumulative incidence can help predict the timing of an epidemic peak under reopening scenarios and the extent of social distancing required to prevent a major surge of new infections in the future.

In order to design a valid seroprevalence study, researchers need to calculate the minimum sample size required to obtain an estimate that is sufficiently precise, requiring an estimate of the proportion of individuals who would test positive for antibodies. Model projections offer the opportunity to evaluate this quantity on future dates during which a proposed seroprevalence study might be conducted. Figure 4 shows projections of cumulative incidence among living individuals who might be sampled and tested in a future seroprevalence survey.

**Figure 4:**
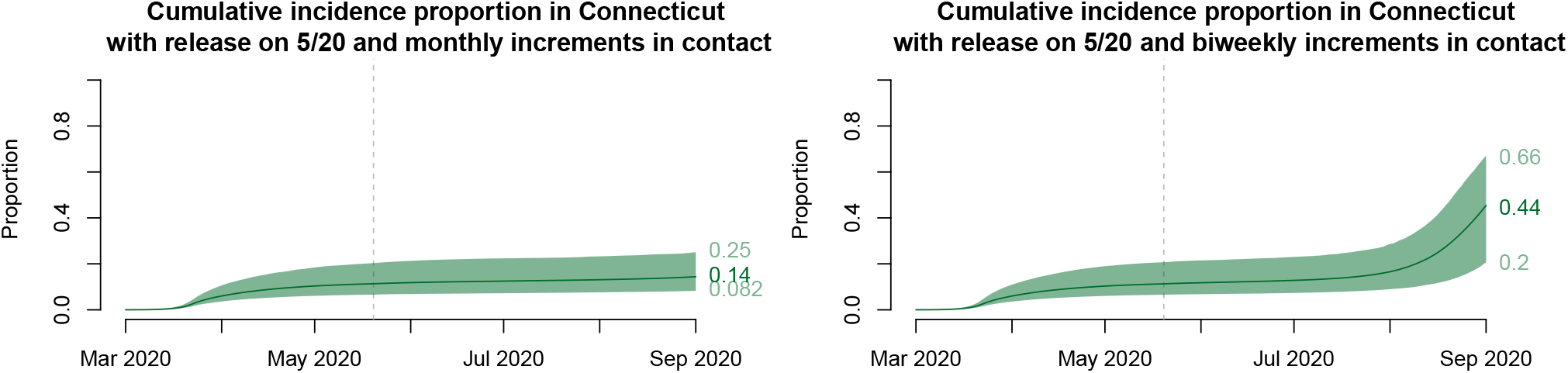
Cumulative incidence proportion among currently alive people under the “slow” (left) and “fast” (right) reopening scenario. Prospective planning of seroprevalence studies requires estimates like these of cumulative incidence estimated at future dates on which sampling might occur. The figure is modified from [28], doi: https://doi.org/10.1101/2020.06.12.20126391.

### Projections beyond Summer 2020

In this report, we have focused on projecting incidence, hospitalizations, and deaths for Summer of 2020. Several factors make long-term projections challenging and potentially unreliable given currently available information about the state’s reopening plans, interventions, and the epidemiology of COVID-19. First, there is uncertainty in how contact patterns may change as a result of state and local policies. As nearby states reopen, and interstate travel increases, outbreaks outside of Connecticut may have a significant impact on incidence within Connecticut. It remains unclear whether schools and universities will reopen according to a standard or altered schedule in the Fall [42, 43]. Second, epidemiologists do not yet know how infectious school-age children may be when they are infected, but it is known that children are unlikely to experience severe symptoms when infected with COVID-19 [23, 44–47]. Epidemiological studies have provided conflicting evidence regarding susceptibility, viral shedding and transmissibility in children [42, 48–50]. Because Connecticut closed K-12 schools approximately only one week before the stay-at-home order took effect, we do not have specific information about the isolated effect of closing schools on the subsequent reduction in transmission. If child care and summer camps reopen as planned at the end of June [51], we may be able to gain information about transmissibility in children from any subsequent increase in cases observed among people who are more likely to be symptomatic. This information will be useful in projecting COVID-19 dynamics in the Fall if K-12 education will take place in person. Third, we do not yet know whether COVID-19 transmission is sensitive to temperature or seasonal changes [34, 52]. If transmission is suppressed over the summer due to warmer temperatures, then Connecticut may enjoy low case counts and hospitalizations over the Summer, with a possible resurgence in the Fall as children return to school and temperatures cool. Another source of long-term uncertainty is related to potentially unequal depletion of susceptible individuals, depending on their risk profile. If a high proportion of older or otherwise high-risk people have already been infected, then we might expect a lower proportion of severe infections in the future, even if overall transmission remains high [53, 54]. Finally, the possibility of improved testing capacity, antibody tests, vaccines, or other therapeutics makes the effect of possible future public health and medical interventions difficult to predict.

## Data Availability

All data and code used in this analysis are available from https://crawford-lab.github.io/covid19_ct/.

## Acknowledgements

We are grateful to Ted Cohen, Robert Heimer, and Edward H. Kaplan, for helpful comments. We thank the Connecticut Hospital Association for providing data on COVID-19 hospitalizations and deaths.

